# Defining Constructs of Hand Function Quality at Home After Cervical Spinal Cord Injury Through Egocentric Videos: A Qualitative Inquiry

**DOI:** 10.64898/2026.11.18.26368413

**Authors:** Adeeba Naoreen, Tanureet Bajwa, Janelle Unger, José Zariffa

**Author notes:** Corresponding author: José Zariffa.

## Abstract

**Introduction:** Loss of hand function is an impactful consequence of cervical spinal cord injury (cSCI), affecting activities of daily living (ADLs). Hand function assessments play an important role in developing interventions that mitigate these impacts. However, assessment can be complicated by factors including the diversity of grasps and use of assistive devices. Consequently, standardized assessments in clinical environments do not fully reflect function in real-world settings. This gap has resulted in increasing interest in assessing hand function at home through wearable cameras (egocentric video) to provide information about real-world hand use. Despite advancements, a framework is required to describe the quality of hand use when observing unconstrained tasks through egocentric video. This study aimed to define constructs that describe hand function quality in unconstrained, in-home activities after cSCI, captured through egocentric videos.

**Methods:** A qualitative study was conducted using focus groups (*N*=17) to generate a list of constructs defining good quality hand use in unconstrained environments after cSCI through egocentric videos. Individuals with cSCI, physicians, therapists, and researchers participated. Content analysis was used to identify patterns, codes and categories.

**Results:** Fourteen categories and thirty-three constructs were identified. While several constructs were common across groups, distinct perspectives also emerged highlighting the multidimensional nature of hand function.

**Discussion:** Perceptions of meaningful hand function encompass a broad range of constructs, emphasizing the importance of both functional performance and motor control aspects. The identified constructs provide a basis for the development of novel metrics of hand function in the context of real-world ADLs.

## Introduction

Individuals with cervical spinal cord injury (cSCI) often have limited hand function, causing difficulties in performing daily activities.^1,2^ There is considerable emphasis on restoring hand function to help improve the quality of life for this population,^2,3^ and measuring hand function is one of the first steps to address deficits, monitor improvement, and modify treatment plans accordingly.

The World Health Organization’s International Classification of Functioning, Disability and Health (ICF) framework is used to measure health and disability through a multidimensional model exploring three domains: body functions and structures, activities, and participation, while considering personal and environmental factors.^4^ Currently, there are various standardized clinical assessments being used to measure hand function, however commonly used assessments primarily address two of the ICF domains: body structures and functions, and activity. Within the activity domain, which explores the execution and limitations of a task by an individual, there are two subdomains: capacity and performance. *Capacity* refers to what an individual can do under ideal or controlled conditions, which is an important first step to implement evidence-based protocols, monitor progress, and evaluate the efficacy of interventions.^5–7^ Capacity is reflected in current outcome measures for upper limb function after cSCI, such as the Graded Redefined Assessment of Strength, Sensibility, and Prehension (GRASSP), the Toronto Rehabilitation Institute Hand Function Test (TRI-HFT), and the Capabilities of Upper Extremity (CUE-T).^8–10^ These clinical tools are typically used in controlled clinical settings assessing isolated activities, such as opening a jar without examining the pre- and post-activities associated with the task. While capacity assesses individuals under controlled settings, the *performance* subdomain refers to what an individual can do in their real-life environments. Current assessment tools are unable to sufficiently capture the diversity of unconstrained, real-world function,^11^ considering the greater variety of tasks, environmental barriers, and the role of caregivers.

The lack of well accepted outcome tools that capture the performance subdomain of the ICF has resulted in increasing interest in observing hand function within the home context using wearable technology including accelerometers, inertial measurement units (IMUs), and egocentric, or first-person, videos.^12–18^ A distinct advantage of egocentric videos compared to other wearables is that they provide both a clear view of the hands as well as contextual information during object manipulation. While egocentric video’s direct data about hand use in the home context is highly valuable, the extended naturalistic observations are more difficult to quantify than standardized outcomes since individuals may perform a wide variety of tasks under diverse environmental conditions. In order to develop metrics that can quantify the quality of hand use in home-based egocentric video, there is a need to first define the constructs that will be measured. The contribution of this study is to describe constructs of hand use quality that can be measured using egocentric video of unconstrained activities at home after cSCI.

## Methods

### Study design

A qualitative study was conducted over six months through semi-structured focus groups informed by the methodologies proposed by Krueger and Casey.^19^ Given the complexity of hand function impairment following cSCI, understanding current clinical tools being used to measure hand function was important in the development of the interview guide for the discussions. A literature review of existing upper extremity assessment tools for spinal cord injury (SCI) was completed, with a review of the ICF to identify key constructs relevant to describing hand function.

### Participants

Purposive sampling was used to recruit focus groups participants from the community, tertiary care hospital sites, and a university in Ontario, Canada. Participants represented diverse backgrounds with experience in hand function and SCI as confirmed through a screening questionnaire. Seventeen individuals were recruited from the following interest groups: individuals living with cSCI, researchers, allied health clinicians, and physicians. All researchers and health care providers (allied health clinicians and physicians) had at least 3 months of experience working with or researching SCI. Individuals with cSCI were 18 years of age or older, had a cervical SCI, and were living in the community with some use of their hands in regular activities of daily living (ADLs). Exclusion criteria for individuals with cSCI included the presence of other neuromuscular disease affecting upper extremity, current admission to the hospital as inpatients, and the inability to communicate in English.

### Procedure

All study procedures were approved by the Research and Ethics Board of the University Health Network (REB#: 24-6060). Prior to the focus groups, participants reviewed five sample two-to-three-minute egocentric videos of individuals with cSCI using their hands in everyday tasks to provide context for the discussion (Figure 1). The egocentric videos were drawn from a previous study.^12^ Participants were categorized into groups based on their role and five 60-minute focus groups were conducted virtually for participant convenience, using a predetermined set of semi-structured questions. Virtual focus groups have been shown to generate a similar level of idea diversity to in-person focus groups.^20^

**Figure 1.**
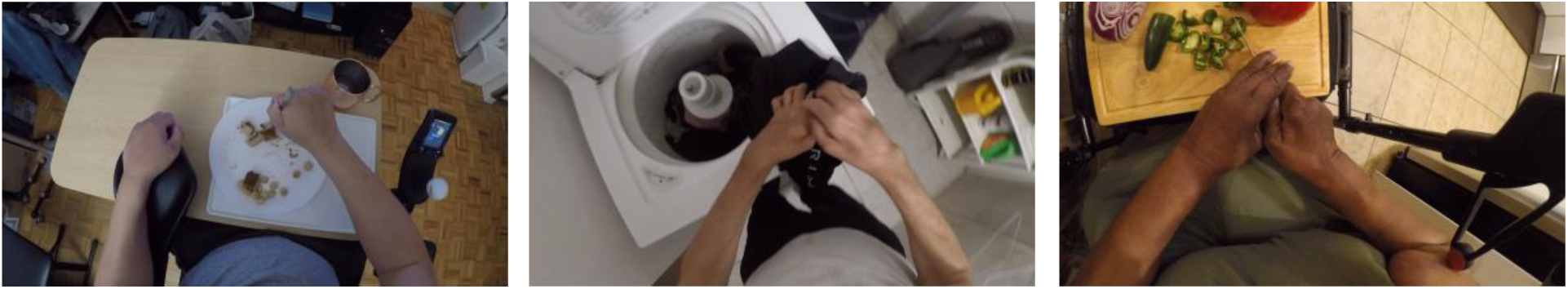
Egocentric video frames capturing individuals with cSCI performing activities of daily living.

Insights from the ICF and commonly used clinical outcome measures for cSCI, including the GRASSP, which provides a detailed and validated evaluation of hand sensorimotor abilities, and the SCIM, which is one of the few assessments to subjectively measure upper limb function in the community after SCI, were used to develop the interview guide.^8,21^ The ICF includes codes which are relevant to real-world hand use, such as self-care and domestic life. To identify appropriate ICF codes, the comprehensive ICF core set for individuals living with chronic SCI was reviewed, including categories that require real-world hand use.^22^ The ICF, the GRASSP, and the SCIM were reviewed to identify important constructs of upper limb function that could facilitate open-ended discussions, while also allowing participants to identify constructs not represented through existing measures.

Participants were asked to share their perspectives on hand function quality in individuals with cSCI and on aspects of hand function that matter most, considering elements of hand function that can be clearly observed by analyzing egocentric video. The fifth focus group suggested data saturation had been achieved, as no new aspects emerged. The complete interview guide can be found in the supplemental material. All five focus groups were conducted in English and moderated by the first author (AN) following details from Kreuger and Casey’s guidelines for focus groups.^19^ Transcript summaries and constructs that emerged from the data were provided to participants for member-checking.

### Data analysis

Data analysis was completed by one graduate student and registered occupational therapist specializing in hand therapy (AN), one occupational therapy student (TB), one independent investigator with a background in physical therapy (JU), and one independent investigator with a background in biomedical engineering (JZ). The research team engaged in ongoing reflexive reflection throughout the research process, mindful of their own positions as clinicians, scientists, and students. Data analysis was conducted following Krippendorff’s content analysis methodologies.^23^ Content analysis was used to systematically identify and organize meaningful perspectives into codes and categories to answer our research question. The focus group were audio recorded, transcribed verbatim, and de-identified. An initial list of quotes was developed through preliminary open coding of five transcripts using NVivo by the first author (AN). Next, all transcripts were organized using conventional content analysis to analyze patterns and systematically code the data by AN and TB.^23,24^ As part of this process, an *a priori* decision was made that constructs unobservable through egocentric videos would not be further explored as they are beyond the scope of this study. Through an iterative process, the codes were rephrased, summarized, condensed, and categorized. The codebook was developed and iteratively reviewed with the principal investigators (JZ and JU).

## Results

Five focus groups were conducted with a total of seventeen participants (Table 1). All individuals with cSCI were from the community, health care providers were recruited from tertiary care hospital sites (including a spinal cord rehabilitation center), and researchers were recruited from the university and the tertiary care hospital sites. Thirty-three observable constructs with fourteen overarching categories emerged from the data, representing perspectives of what experts considered indicative of good hand function quality in people with cSCI through egocentric videos. At least one participant from each focus group provided feedback in agreement with the results during the member-checking process. The complete codes and categories are presented in Table 2, with the summative distribution of the codes presented in Table 3.

**Table 1.** Participant characteristics (*N*=17)

|  |  |
|---|---|
| Sex, female, *n* (%) | 9 (53%) |
| Occupation, *n* (%) |  |
| Individuals with SCI | 4 (23%) |
| Health Care Providers |  |
| Physical Therapist | 2 (12%) |
| Occupational Therapist | 2 (12%) |
| Physician | 4 (23%) |
| Researchers | 5 (29%) |

**Table 2.** Categories and codes identified in the content analysis.

| Categories | Codes | Representative quotes |
|---|---|---|
| Time | Performing tasks in a timely manner | *Time is major, because most of us have appointments, right? So, we have to get to our appointments on time, and we have to make sure that we're actually downstairs in order for Wheel-Trans to not leave us.* (P10, individual with cSCI) |
|  | Speed of hand movements | *Like sometimes, if they're doing it, they could be doing it too fast, so we don't want them to do it that fast. We need you to slow down so we can get better quality.* (P01, allied health clinician) |
|  | Task efficiency | *It was interesting to see people set up and try things a couple of times. And ideally, if you got to good hand function, it would be one time and effective, right?* (P14, physician) |
| Task completion | The ability to complete a task | *I think when I think about function, I always just think about getting it done, like functionally, what I can do to achieve the task.* (P02, allied health clinician) |
|  | Being able to successfully set up and clean up the task | *So just thinking about, is it the act of the cutting that's the task or is it actually be able to do it from beginning to end? So to me, I think good hand function looks like being able to do it from beginning to end.* (P14, physician) |
|  | Ability to perform activities of daily living | *Good hand function would mean that you're able to get yourself dressed, you're able to help yourself eat. You're able to handle your hygiene. All of those things come from good hand function. And if you don't have good hand function, then those vital things that you need to complete and do, how would you be able to do them?* (P10, individual with cSCI) |
|  | Task sequencing | *Doing many, many, many steps before getting the tube open. So it's like, it's not only time, it's only, also how many things that person needs to do before getting the task done.* (P16, physician) |
| Control and coordination | Fine motor control | *So if they're getting it done but we actually think they could do it with more dexterity, then it wouldn't be good hand function, that wouldn't be good quality for that person.* (P01, allied health clinician) |
|  | Fluidity of movements | *His intentions were there, and he was able to follow through with his intentions, versus chasing something to the corner, like the first person trying to open the Advil over there, chasing it to the corner and trying to cut it, holding it in the corner. All ingenious ways of accomplishing, but that wasn't very fluid, in control, or strength-orientated by any means.* (P13, individual with cSCI) |
|  | Ability to manipulate objects | *So, they are constantly trying to change the contact points in order to make a manipulation. How much they're efforting to change this contact point to find a stable contact point to make the manipulation is something that I, that I care a lot […].* (P08, researcher) |
|  | Ability to hold objects | *To hold the bottle and grip the bottle, and then use my right hand to open. That was a big, big accomplishment for me.* (P12, individual with cSCI) |
|  | Precision of hand use | *Usually people with better control, their movements are observably more precise.* (P17, physician) |
| Safety | The ability to use hands safely | *I was also very afraid when the person grabbed a knife, right, and started using it, so I would also say, you know, it's probably a good indicator of safety.* (P07, researcher) |
| Range of motion | Mobility of hands and fingers | *I would grade with range of motion of first the thumb. Do you have […] partial range or full range […].* (P11, individual with cSCI) |
| Consistency | Repetition of tasks | *So that potentially would be useful if they can consistently demonstrate the same movement more than once.* (P17, physician) |
|  | Fluctuations in hand performance | *I was interested in how the performance fluctuated across the task.* (P14, physician) |
| Hand grasp variety | The variety of hand grasps | *But on the overall spectrum of things, the more variations of grasps you can achieve, you know ideally the more number of activities you can achieve in turn.* (P02, allied health clinician) |
| Breaks | Pauses during hand use | *You're willing to do it, but you also have to pause, because you can't do it like a normal person.* (P12, individual with cSCI) |
| Independence | The ability to perform tasks independently | *They want to be able to do by themselves toileting. That's their dignity.* (P05, researcher) |
|  | The ability to perform tasks independently with an assistive device | *If you're struggling with your fork, you might pick up some food, and then now your fork becomes heavier because of what you put it on there, but a strap support for the hand and the thumb then might make it easier for you to do it, right?* (P10, individual with cSCI) |
| Assistive devices | Appropriate use of assistive devices | *Well, you know, ideally with devices, can the person don it themselves? Or like donning and doffing independently or not, that would be another part of that.* (P14, physician) |
| Task complexity | Variety of tasks performed in activities of daily living | *If you are using a wide variety of the grasps, meaning that you are performing different activities, you are independent, you have […] a higher clinical matrix.* (P08, researcher) |
|  | Level of task difficulty | *I guess I would say if they're able to do more difficult tasks, that would probably reflect the overall better hand function.* (P17, physician) |
| Compensation | Using compensatory strategies | *And you know it can be hands and teeth or hands and a door jam, you know. Like what we saw in the videos […] Or just whatever strategy, you know, it could be environment and equipment that's available to help.* (P04, allied health clinician) |
|  | Using modified and/or adaptive grasps | *Your hand will not be the same as before your stroke or your spinal cord injury. But your body, the pathways, gonna find a way to strategize the movements. You gonna hold the spoon maybe differently […]* (P05, researcher) |
|  | The use of environmental resources | *I mean, it's clear that the person leans against the wall, like he secures the object against the wall, to be able to achieve that particular function, okay? So, for that particular task. So that's important.* (P07, researcher) |
|  | Environmental modifications to support task | *I wondered if his control got worse reaching further away?* (P04, allied health clinician) |
| Errors | The number of errors during a task | *If you're cleaning up, picking up a pencil, I wouldn't be too upset if you had to do two tries. If you're trying to raise a cup to your mouth and you're missing and burning yourself, that's a whole different…* (P14, physician) |
|  | Dropping items during task | *If you're losing an object, then obviously there needs to be an improvement in your ability to do whatever task it is that you have at hand.* (P09, researcher) |
| Bimanual vs unimanual hand use | Performing bimanual tasks | *I could not lift my left and rub my… now, when I wash my face in the shower, it's like I try to use both hands. I remember, the 500ml water bottles, I used to hold it with my right hand, and then use my teeth to, to open it. Now, I notice that […] I'd grip it with my left hand, and use the right hand to open it […]* (P12, individual with cSCI) |
|  | Performing unimanual tasks | *But if you are only looking for quality of hand function, the patient that can do the task with one hand, of course, is a better quality, I think, is a better quality of hand function than do it with two hands.* (P16, physician) |
|  | Using bimanual compensation | *Typically, people will not use two hands to improve the quality of the movement. They will use it more to be able to complete the task that they want to complete, right?* (P09, researcher) |
|  | Bimanual coordination | *I think it had something to do with actually using the other hand to help as opposed to trying to do everything just with the one hand.* (P15, physician) |

**Table 3.** Summative frequencies of codes.

| CODES | Allied health clinicians | Researchers | Individuals with SCI | Physicians |
|---|---|---|---|---|
| Performing tasks in a timely manner | 7 | 6 | 5 | 10 |
| Speed of hand movements | 3 | 3 |  |  |
| Task efficiency | 2 | 1 |  | 5 |
| The ability to complete a task | 6 | 4 | 7 | 3 |
| Being able to successfully set up and clean up the task |  |  |  | 2 |
| Ability to perform activities of daily living | 3 | 3 | 1 | 3 |
| Task sequencing | 3 |  |  | 3 |
| Fine motor control |  | 1 | 6 |  |
| Fluidity of movements | 1 |  | 2 | 4 |
| Ability to manipulate objects |  | 2 | 2 |  |
| Ability to hold objects | 2 | 5 | 4 | 2 |
| Precision of hand use |  | 1 |  | 6 |
| The ability to use hands safely | 6 | 3 | 6 | 7 |
| Mobility of hands and fingers | 1 | 3 | 7 | 4 |
| Repetition of tasks |  |  |  | 6 |
| Fluctuations in hand performance | 1 |  |  | 5 |
| The variety of hand grasps | 6 | 7 | 1 | 6 |
| Pauses during hand use | 2 |  | 4 | 2 |
| The ability to perform tasks independently | 3 | 7 | 2 | 6 |
| The ability to perform tasks independently with an assistive device | 3 |  | 2 | 3 |
| Appropriate use of assistive devices | 4 |  | 1 | 12 |
| Variety of tasks performed in activities of daily living | 3 | 4 |  | 2 |
| Level of task difficulty | 1 |  |  | 3 |
| Using compensatory strategies | 6 | 2 | 4 | 6 |
| Using modified and/or adaptive grasps | 5 | 6 | 1 | 5 |
| The use of environmental resources | 4 | 3 | 1 | 1 |
| Environmental modifications to support task | 3 |  |  | 1 |
| The number of errors during a task | 1 |  |  | 2 |
| Dropping items during task | 3 | 1 | 2 | 4 |
| Performing bimanual tasks | 1 |  | 2 | 2 |
| Performing unimanual tasks |  |  | 2 | 2 |
| Using bimanual compensation | 1 | 4 | 6 | 2 |
| Bimanual coordination | 4 |  | 5 | 7 |

### 1. Time

Three constructs were found relating to time: performing tasks in a timely manner, speed of hand movements and task efficiency. Within this category, performing tasks in a timely manner was a construct which emerged across all groups. Health care providers highlighted that performing tasks in a timely manner is important as this is a determining factor to organizing the day and when “*you consider life with a spinal cord injury, everything takes so much longer, and people are going to get to a point where they just want to get stuff done.”* (P04, allied health clinician). Allied health clinicians and researchers shared that beyond assessing time for completing a task in determining quality, it is important to consider the speed of hand movements because “*where going slow and really, taking your time with activity yields more success, but other times when people use momentum, they get more success with that as well.”* (P02, allied health clinician). A unique perspective regarding time mentioned efficiency; “*ideally, if you got to good hand function, it would be one time and effective, right?”* (P14, physician).

### 2. Task completion

Task completion includes the ability to complete a task, being able to successfully set up and clean up the task, ability to perform activities of daily living, and task sequencing. The ability to complete a task was an important construct that was discussed throughout all focus groups, highlighting *“he was able to figure it out. That’s what I got from it. It might not look as we want it to look 100%, but he was eating, and he figured out how to eat his meal.”* (P10, individual with cSCI). A unique perspective reported that the ability “*to get the tools that you need and return them”* (P14, physician) should be observed rather than observing the task in isolation.

### 3. Control and coordination

There were five constructs under control and coordination including fine motor control, fluidity of movements, ability to manipulate objects, ability to hold objects and precision of hand use. Participants shared that being able to hold objects functionally was important, for example, noting as an improvement that they valued “*I needed something around my hand to help me hold it […] now it’s gone, now I don’t need it”* (P10, individual with cSCI).

### 4. Safety

The ability to use hands safely emerged from all groups particularly due to the potential for impaired sensory function and *“If you don’t feel the pain, or you’re not… you don’t know you’re injured, then it be… it can be detrimental to your health, right?”* (P12, individual with cSCI).

### 5. Range of motion

Range of motion includes the mobility of hands and fingers, with participants sharing *“good hand functions is to have mobile fingers and thumb, and to be able to control it. Maybe not as well as normal. […] but we need mobile fingers.”* (P11, individual with cSCI). If an individual has limited movement where “*He only moved his thumb, that’s not a good hand function.”* (P11, individual with cSCI). This is important because *“if you don’t have great range of motion, of course, you can’t grasp things.”* (P16, physician).

### 6. Consistency

Two constructs were identified under consistency including the ability to repeat tasks, and fluctuations in hand performance throughout a task. These constructs were not discussed across all groups but were emphasized particularly in both the physician groups. Participants shared that

*“seeing the repetition of a fluid task, being able to do something repetitively well to me would be a sign of quality”* (P14, physician). Beyond repetition, participants noted the importance of demonstrating the same level of hand function across the task in determining quality as “*consistency comes to mind*” (P02, allied health clinician) and the variability and “*how consistently they can do a movement as far as control and coordination*” (P17, physician) as important.

### 7. Hand grasp variety

Hand grasp variety refers to the different types of grasps an individual can use. This was suggested *“because we don’t just eat; we dress, we call the phone, we hold the phone […]”* (P05, researcher). Participants emphasized it’s important to consider because by *“looking at the types of grasps and how many they can successfully achieve kind of opens up the amount of you know, activities they can do”* (P02, allied health clinician). Examining grasp types was also suggested as a subtle variable linked to independence, because *“if we just track the hand grasp, and to track the grasp they are using, we are able to infer that, are they independent or are they using a wide variety of tasks?”* (P08, researcher).

### 8. Breaks

Pausing during hand use was a general construct that emerged from all groups except for researchers. Participants identified that pauses are not only important, but *“breaks were mandatory. If I didn’t take the break, that I wouldn’t even be able to complete the task*.” (P10, individual with cSCI). The physician group shared that in order to “*have a precise task or movement, probably that person needs to take more pauses just to be precise.”* (P16, physician).

### 9. Independence

Individuals with cSCI shared their experience that learning to be independent is essential because “*what if there’s nobody there to help?”* (P10, individual with cSCI), and “*if you don’t have good hand functions, your independence is compromised*” (P12, individual with cSCI). This included being independent with the use of an assistive device because *“if you rate as critical being independent, of course, hand function with device or adapted grasp is great because you are achieving independence”* (P16, physician).

### 10. Assistive devices

Perspectives differed across and among groups regarding the use of assistive devices during tasks as a component of hand function quality. Participants suggested *“there could be better function through some other assistive, with like modified devices*” (P03, allied health clinician), and if *“using aids appropriately, that would be a sign of good hand function*.” (P14, physician). It was expressed that using assistive devices could demonstrate poor function because *“If we don’t have good hand functions, we need tools to assist, even though tools might not be great”* but highlighted that *“it’s better than using your teeth.”* (P11, individual with cSCI). Similarly, another participant conveyed that *“If they’ve adapted and are using other modalities of doing the task without actually using their hands or without, then they don’t have good hand function. They’ve just learned how to do a task*.” (P15, physician).

### 11. Task complexity

Allied health clinicians and physicians communicated an intersection between being able to perform a variety of tasks and being able to assume a variety of hand grasps, and *“if you can do…like a more variety, a broader variety of tasks, it means that you have better hand function.”* (P16, physician).

### 12. Compensation

Participants emphasized that because *“hand function is very key to independence”,* that using compensatory strategies is appropriate such as *“hands and teeth […]”* (P04, allied health clinician). The use of adaptive grasps was discussed among all groups and that *“it’s okay if it’s in a different manner, you know, kind of, to open a bottle or something”* (P04, allied health clinician). Researchers appreciated that after a cSCI “*your body, the pathways, gonna find a way to strategize the movements*”, and using new and different grasps is acceptable as it “*is their way. […] you are the boss of your own body.”* (P05, researcher). Physicians highlighted that using adaptive grasps are acceptable if they are appropriately posturing the movements, regardless of level of injury. However, inappropriate posturing would be deemed as poor quality function. A caveat to this perspective was identified when comparing two individuals with cSCI, in which an individual with a less severe injury completing a task without an adaptive grasp would be rated higher than an individual with a more severe injury completing a task with modified grasp.

Regarding environmental supports, participants described *“Whether it be the door, putting it in the door, or whatever else. […] All of us can agree with that, that when you have this injury, you’re forced to figure it out. To explore different types of ways that you can try to complete a task.”* (P10, individual with cSCI), and *“when the environment becomes an aid, I think that that should be important.”* (P07, researcher).

### 13. Errors

The health care provider groups indicated “*errors and number of errors, they make while performing the task*” (P02, allied health clinician) as an important perspective, as well as whether “*[they] can achieve what they want to do without complications.*” (P16, physician). More specifically, dropping items as an indicator of poor-quality hand function was discussed among the groups, and that it *“does factor to how you assess the quality of their movement not only doing the task, but doing the task without any complications such as spilling stuff all over themselves.”* (P17, physician).

### 14. Bimanual versus unimanual hand use

The ability to use hands cohesively was regarded as important in assessing the quality of function, with an individual with cSCI sharing *“I remember, the 500ml water bottles, I used to hold it with my right hand, and then use my teeth […] to open it. Now, I notice that I’d grip it with my left hand, and use the right hand to open it.”* (P12, individual with cSCI). Researcher shared differing opinions on using bimanual compensation, with some supporting the use of the contralateral hand to compensate for the ability to complete a task with one hand, *“I don’t see any problem whatsoever in using both hands for what would be a typically, unilateral hand, if that is the focus*.” (P07, researcher). Whereas others disagreed that *“if you’re using both hands to complete a task that ideally, which an able-bodied person would do using just one hand, then I want to say that the idea is completion of task […]”* because *“Typically, people will not use two hands to improve the quality of the movement.”* (P09, researcher).

## Discussion

The focus groups revealed perspectives from experts including individuals with cSCI, researchers, allied health clinicians, and physicians on how to define the quality of hand function among individuals with cSCI performing unconstrained ADLs in their home environments, as assessed using egocentric video. The distribution of codes across the groups demonstrated some similarities in perspectives such as performing tasks in a timely manner, ability to complete a task, ability to use hands safely, variety of hand grasps, ability to perform tasks independently, using modified and/or adaptive grasps, dropping items during task, and using bimanual compensation. However, there were also differences; certain constructs were emphasized within specific groups suggesting that hand function quality can be influenced by an individual’s lived experience, clinical role, or research perspective. Health care providers identified several constructs which were not explored in other groups such as task difficulty, sequence of actions, fluctuations in performance, and errors during a task, whereas individuals with cSCI more frequently highlighted constructs such as fine motor control, the ability to move their fingers and hands, using bimanual compensation, and the ability to control and hold objects.

Hand use assessments provide a reliable way to evaluate hand function and assist in supporting clinical decisions. There were several constructs identified in this study that are consistent with current literature, such as performing tasks in a timely manner. Time is commonly assessed in upper extremity outcomes such as the Jebsen Hand Function Test, and the CUE-T.^10,25^ Independence is another construct that is evident in the literature and outcome tools, such as the SCIM, as one of the most important goals in SCI rehabilitation.^26^ Another finding in line with evidence from currently used outcomes measures is errors during task performance. This construct suggests that although a person may be able to complete a task, frequent errors would demonstrate poor quality in their function. Assessments such as the Van Lieshout test (VLT), the TRI-HFT, and the GRASSP either measure or document errors during assessment, which can impact an individual’s overall score.^9,27,28^ For example, the prehension components of the GRASSP examines several of the constructs identified such as numbers of drops, the ability to produce specific grasp patterns, and dexterity in fine motor tasks.

There were differing opinions on whether using assistive devices is evidence of good hand function quality. Literature has shown that assistive technologies enable individuals with disabilities to live productive and independent lives^29^ and can improve upper limb function after SCI.^30^ In the physician group, some participants noted that if an individual is using adaptations or other modalities, it does not necessarily reflect quality hand function but rather that they have learned alternate strategies to perform the task. A physician’s role may lead them to focus on hand function from a motor capacity perspective,^31^ thus, the reliance on equipment may be an indication that the individual is unable to perform the task using their own hands or are limited in their function. Allied health clinicians, on the other hand, may view tools differently as they place greater emphasis on functional ability, meaningful participation, and independence, where tools could be an asset.^32^

The focus groups showed task completion to be a consistent and important component of hand function quality. Although task completion is observed through outcome measures such as the GRASSP, SCIM, TRI-HFT, and Sollerman Hand Function Test, it is often measured in conjunction with grasp quality and time. Participants may have heavily emphasized task completion as it is a meaningful and relevant way to evaluate how well someone can use their hands in everyday life. As noted by individuals with SCI, support at home may be limited, and from their perspectives, regaining the ability to participate successfully in daily activities is integral in reintegrating into their regular routines and independence. Individuals with cSCI did not report precision of hand use as important to measuring quality, which may be due to prioritizing task completion over how well the task is completed, and often precision may require excessive time and not be practical. Allied health clinicians strongly emphasized that being able to complete a task with adaptive strategies is more important than being able to emulate typical grasp types. Health care providers and researchers acknowledged that functional performance can be influenced by an individual’s physical ability, the environment, and task demands, such as using environmental modifications, distance, or advanced preparation, and should be considered.

Health care providers shared that it is important to establish what is appropriate for an individual’s level and type of injury. For example, the same function cannot be assumed for a person with a complete and incomplete injury. Creating normative values for different levels and severities of injury is important for establishing expected functional abilities. However, considering that comparison with normative values would require a person’s medical history and injury information, performance relative to expectations was not included as a construct observable from egocentric video alone.

Notably, several constructs may also serve as proxies for underlying concepts that are important to hand function but cannot be directly observed through egocentric video. Often health care providers assess sensory function along with motor functions, and although sensation is not an observable construct through egocentric videos, it could generally be captured indirectly through measurable constructs such as safety or errors by observing repeated drops, or contact with a sharp, hot, or harmful object without a reaction. Similarly, other unobservable constructs were expressed such as confidence, persistence, and strength, and although they cannot be directly measured through video, they could be indirectly assessed through observable constructs such as task completion, the ability to hold an object, dropping items, fluctuations in hand movement, and using bimanual compensation.

### Limitations

Although this study reached data saturation, it was limited in the number of participants recruited from an isolated geographical region. Participants were recruited from Toronto, Canada, and results therefore may not be generalizable to a wider population. However, diverse perspectives were included to provide a comprehensive representation of the various aspects describing hand function quality. We purposively selected participants to achieve a broad and experienced panel to understand the research question from a rehabilitation, research and personal perspective.

## Conclusion

We explored perspectives from multiple groups to define constructs of hand function quality in unconstrained activities after cSCI, as observable through egocentric videos. Through the focus groups interviews, we identified thirty-three constructs in fourteen categories, with differing emphasis across participant groups. While some redundancies and overlap remain among the thirty-three constructs, these concepts were intentionally retained to comprehensively reflect the focus group discussions. This study provides a foundation for measuring the quality of hand use during of performance of real-world ADLs. Findings will assist in evaluating the efficacy of interventions and monitoring progress to assist with recovery of hand function. Future work will include recruiting a larger panel of collaborators to reduce the list of constructs into measurable metrics that can be implemented to evaluate hand function quality through egocentric videos.

## Acknowledgments

The authors would like to express their sincere gratitude to all participants for generously sharing their time and insights to this research, and Sharmini Atputharaj for leading participant recruitment.

## Author contributions

AN: Methodology, formal analysis, investigation, writing – original draft preparation, funding acquisition, project administration. TB: Formal analysis, writing – review & editing. JU: Conceptualization, methodology, formal analysis, writing – review & editing, funding acquisition, project administration, supervision. JZ: Conceptualization, methodology, formal analysis, writing – review & editing, funding acquisition, project administration, supervision.

## Statements and declarations

### Ethical considerations

This study was reviewed and approved by the Research Ethics Board (REB) of the University Health Network (UHN) (REB #24-6060).

### Consent to participate

All subjects signed written informed consents.

### Consent for publication

Not applicable.

### Data availability

The data analyzed during the current study are available upon reasonable request to the authors and completion of a data sharing agreement.

## Supplementary Material

### Focus Group Interview Guide

1. **Outside of a specific clinical assessment, what does “good hand function” mean to you?**

Prompt: How important is the ability to perform daily activities independently, such as dressing, eating, and grooming, in determining good hand function and why?

**2. In the videos that you have watched at home, what did you conclude about the person’s hand function and why?**

Prompt: What were some of the observations that stood out to you regarding hand use? Prompt: What were some strengths and weaknesses of the videos to evaluate hand function based on your observations?

**3. How can observing a variety of grasps an individual uses successfully (power, lateral, pinch) be effective in determining the quality of function?**

Prompt: Why may or may not observing more variety indicate better hand function?

**4. What aspects of hand coordination during a bilateral task, like opening a water bottle, might be most useful to observe, and why?**

Prompt: Does using both hands for a unilateral task (such as using two hands when using a spoon) affect the quality of hand function in individuals with SCI?

**5. How would you take into consideration speed, fluidity, control and coordination when evaluating someone’s hand function?**

Prompt: How might frequent pauses affect the quality of hand function?

Prompt: How might tremors, jerky movements, or dropping items affect the quality of hand function?

**6. How would you take into consideration the ability to adapt or compensate for hand function deficits such as using an adapted grasp or the use of assistive device?**

Prompt: What factors would indicate a successful or less successful adaptation?

